# Representation of the hierarchical and functional structure of an ambulatory network of medical consultations through social network analysis, with an emphasis on the role of medical specialties

**DOI:** 10.1101/2023.08.14.23294067

**Authors:** Fernando Martín Biscione, Juliano Domingues da Silva

**Author notes:** Corresponding author: Fernando Martín Biscione (FMB).

## Abstract

**Background:** Ambulatory Health Care Networks (Amb-HCN) emerge when doctors establish circuits of patient referral and counter-referral in their offices, explicitly or spontaneously. We aimed to characterize the structural and functional topology of an Amb-HCN of a private health insurance provider (PHIP) using objective metrics from graph theory.

**Methods:** A Social Network Analysis was conducted with administrative claim data of a Brazilian PHIP. Included were beneficiaries of a healthcare plan not restricting the location or physician caring for the patient. A directional and weighted network was constructed, where doctors were vertices and patient referrals between doctors were edges. Vertex-level measures were calculated and grouped into three theoretical constructs: patient follow-up; relationship with authorities; and centrality profiles. To characterize physicians into these profiles, cluster analysis was conducted using the non-hierarchical K-means technique.

**Findings:** Between 04/01/2021 and 05/15/2022, 666,263 individuals performed 3,863,222 office visits with 4,554 physicians. Non-primary-care medical specialties (e.g., cardiology, endocrinology etc.) were associated with central profile in the graph, while surgical specialties predominated in the periphery, along with pediatrics. Only pediatrics was associated with strong and prevalent patient follow-up. Weak and shared patient follow-up was present for many doctors from internal medicine and family medicine. All profiles exhibited pairwise relationships with each other, and with clinical characteristics and outcomes of the patients they treated. For example, physicians identified as authorities were frequently central and treated patients with more comorbidities. Eleven medical communities were identified with clear territorial and medical specialty segregation.

**Conclusions:** Viewing the Amb-HCN as a social network provided emerging insights into the most influential actors and specialties, potential gaps in care, and the most prevalent diseases in our patient portfolio. Identifying self-constituted Amb-HCN can form a rational basis for developing more formal networks or monitoring patient care performance without assigning responsibility to single physicians.

## Introduction

Health Care Networks (HCN) can be defined as the way in which health systems organize their health actions and services, in an integrated, functional, and hierarchical manner, according to the different technological densities each one offers, in order to ensure care for the population being served [1]. The operational structure of HCN is therefore constituted by the different actors of health care and the connections that link them. HCN are characterized by the development of horizontal and vertical relationships between the various multi-professional care points for the patient, in a more or less regulated manner. This network organizational arrangement of health systems is justified by fundamentals such as economies of scale, regionalization (or territorial coverage) of care, and guaranteeing quality, sufficiency, and access to health for the population being served [1]. Another basic principle of HCN is the structured levels of care according to technological density and rational use of resources, ranging from the level of lowest density (Primary Health Care) to intermediate density (Secondary Health Care), to the highest technological density level (Tertiary Health Care). It is the responsibility of Primary Health Care to be the first level of care, with a resolution function for the vast majority of the population’s health problems, from which specialized care is activated [1].

Regarding the ambulatory level of health care, this study will conceptualize the ambulatory care network (Amb-HCN) as the circuits of referral and counter-referral established, explicitly or spontaneously, between the doctors who attend to patients in their offices. It is in the context of the doctor’s office where the primary level of health care takes place, with low technological density, and part of the secondary care (specialized consultations). Therefore, characterizing the organic and hierarchical functioning of an HCN through objective metrics can be strategically important for health managers, allowing them to: identify informal patterns or hierarchies among health actors that reveal the forces governing the organic functioning of the network; compare the observed structure of the HCN with the structure of other external networks, the same network over time, or with that expected according to the health care model proposed by the manager for the network under their responsibility; identify actors with positive or negative influence on the HCN, according to the objectives defined by the manager; seek correlations between HCN performance metrics and attributes of health outcomes, quality or value delivered to users; propose changes to reimbursement models based on results, quality and value in health. The ultimate goal will always be to provide health managers with information that enables corrective or preventive decision-making towards continuous improvement of HCN performance.

Characterizing the properties of a Amb-HCN through objective metrics is a complex methodological challenge. Recently, Social Network Analysis (SNA) has received strong interest from the scientific community for the study of numerous health phenomena that are inherently relational, complex, and dynamic, including, but not limited to, identifying relationships and personas, dissemination of innovations, and studying patterns of information exchange or collaboration among actors in diverse areas such as education, health promotion, infectious diseases spread, digital health, management, regulation, etc. [2, 3]. SNA is a set of methods and concepts based on graph theory that analyze systems whose properties stem from the relationship between entities. The value of SNA in determining the properties of HCN has been tested in recent studies. Researchers have used SNA on administrative data to identify hidden or informal referral and counter-referral networks among doctors who treat common patients [4, 5]. By applying SNA on networks of professional teams who care for diabetic patients, Ostovari et al. [6] identified key professionals and healthcare providers in the network. The same researchers found that when primary care physicians had high values in community-level centrality measures (i.e., closeness, betweenness, and degree), the diabetic, hypertensive, or dyslipidemic patients they cared for had lower hospitalization and emergency department visit rates [7]. Similar results were reported in another study, in which patients with cardiovascular diseases who were cared for by healthcare teams with dense interactions and low centralization had 38% fewer hospitalization days and lower healthcare costs compared to patients cared for by teams with less dense interactions that revolved around a few central professionals. Face-to-face dense interactions among team members were also associated with more effective control of hypercholesterolemia and a 73% lower need for emergency department visits [8]. Although these and other studies point to promising results in the use of SNA for analyzing complex HCN, the most appropriate set of measures and evaluation metrics, as well as their clinical and administrative/managerial significance, remains uncertain [2, 9, 10].

The purpose of this study is to characterize the structural and functional topology of the Amb-HCN of a private health insurance provider (PHIP) through objective measures and metrics, based on the referral and counter-referral circuits, whether explicit or spontaneous, established between network physicians during patient care in their offices. Therefore, the study will focus on the role of physicians as the main responsible for generating these circuits and will propose the creation of metrics for the operational definition of Amb-HCN attributes considered important for PHIP strategy. The measures and metrics will be analyzed according to the physician’s specialty, seeking to determine their contribution to the functioning of Amb-HCN, and their relationships with patient clinical characteristics and outcomes will be analyzed.

## Materials and methods

### Study design

This is a cross-sectional exploratory and explanatory quantitative study, with a secondary data analysis study design.

### Setting and period

The study was conducted on the beneficiaries base of a PHIP located in Belo Horizonte, capital city of Minas Gerais state, Southeastern Brazil. This company has a coverage area in Belo Horizonte and 33 other municipalities in its metropolitan region. As of April 2023, it provided assistance to more than 1.538 million beneficiaries and had over 5,300 accredited doctors.

The study considered a base of 1,042,654 beneficiaries who, between April 2021 and March 2022, were beneficiaries of a healthcare plan that did not restrict the location or physician who cares for the patient, remaining at his or her discretion and convenience. The study evaluated all office visits made by patients between 04/01/2021 and 05/15/2022.

### Data and definitions

The study’s database was extracted from secondary databases maintained by the PHIP in its own Data Warehouse, and included the following data: physician identification; physician age and specialty; patient satisfaction ratings reported by the patient after the consultation, on a scale of 0 to 10; patient identification; office medical visits made by the patient, including location, physician who performed the consultation, and date of consultation; number of chronic comorbidities of the patient recorded in the PHIP, classified according to Elixhauser et al., Charlson-Deyo et al., and Feudtner et al. [11, 12]; cost of ambulatory exams and therapies ordered by the physician; number of emergency department visits, hospital admissions due to medical condition, and total number of days of hospitalization due to medical condition of the patient in the same period of the study. All these data represent administrative claim data or beneficiary registration information routinely collected by the PHIP.

The data reported in this research was extracted in April 2023. However, programming codes and methods were developed for business – rather than research – purposes on a subset of the same dataset available to the lead researcher in October 2022, in accordance with the PHIP’s institutional compliance and legal standards. The data was retrieved from data repositories at the individual level in a pseudonymized form and handled anonymously thereafter.

### Data analysis

SNA was conducted to evaluate the properties of the Amb-HCN, conceptualizing the latter as the set of referral and counter-referral circuits of patients established between physicians, either explicitly or spontaneously, who treat patients in their offices. The SNA design followed general principles recommended in Blanchet et al. [13] and De Brún et al. [14]. According to the classification proposed by Benhiba et al. [15], it is a structural SNA analysis (i.e., describing, at discrete intervals, the topology of the network, the roles of the vertices, describing communities and subgroups, etc.) with an egocentric view (i.e., characterizing actors according to the relationship they have with their immediate network). The constitutive elements of this network were as follows:

a) vertices (V): represented by the physicians v*i*… v*j* who performed the consultations in their offices;

b) edges (E): represented by patients who, after a consultation with a particular physician v*i*, had a consultation with another distinct physician v*j* within an interval of 7 to 45 days, thus linking physician v*i* to physician v*j*. This interval was chosen to represent the most likely period in which referrals between professionals occur and would reveal referrals motivated by the same health-related problems;

c) vertex weight (Vw): since the contribution of each physician to the total number of consultations in the network depends on their own characteristics and, at the same time, the specialty to which they belong, the weight of the vertices was represented by the product below:

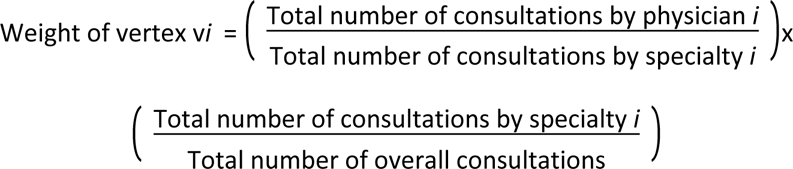

Or

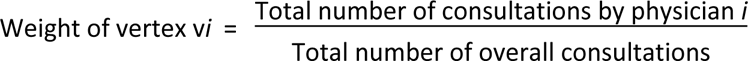

d) edge weight (Ew) between v*i* -> v*j*: represented by the ratio between the number of referrals from doctor v*i* to doctor v*j* and the total number of patients attended by doctor v*i*:

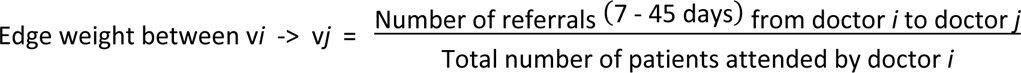

The network thus designed can be understood as a directed and weighted network. It is worth mentioning that, within this health system, the Amb-HCN has a basically self-regulated design, depending on patient characteristics (such as place of residence, personal preferences, etc.), physician characteristics (such as specialty, location of practice, private network of collaboration and trust with other professionals, etc.), and terrain (such as availability of transportation, etc.). There are no referral flows between physicians explicitly promoted by the PHIP.

Several performance metrics were calculated at both the network and vertex levels. For the network, density, diameter, radius, average path length, global efficiency, clustering coefficient, and number of weak and strong components were calculated [16]. The calculation of the weighted versions of the metrics was prioritized, assigning the edges the weight Ew, as previously described. Unweighted versions were also calculated for some metrics for descriptive purposes or when the calculation of the weighted version was not applicable (Box 1).

##### Box 1. Network-level performance metrics

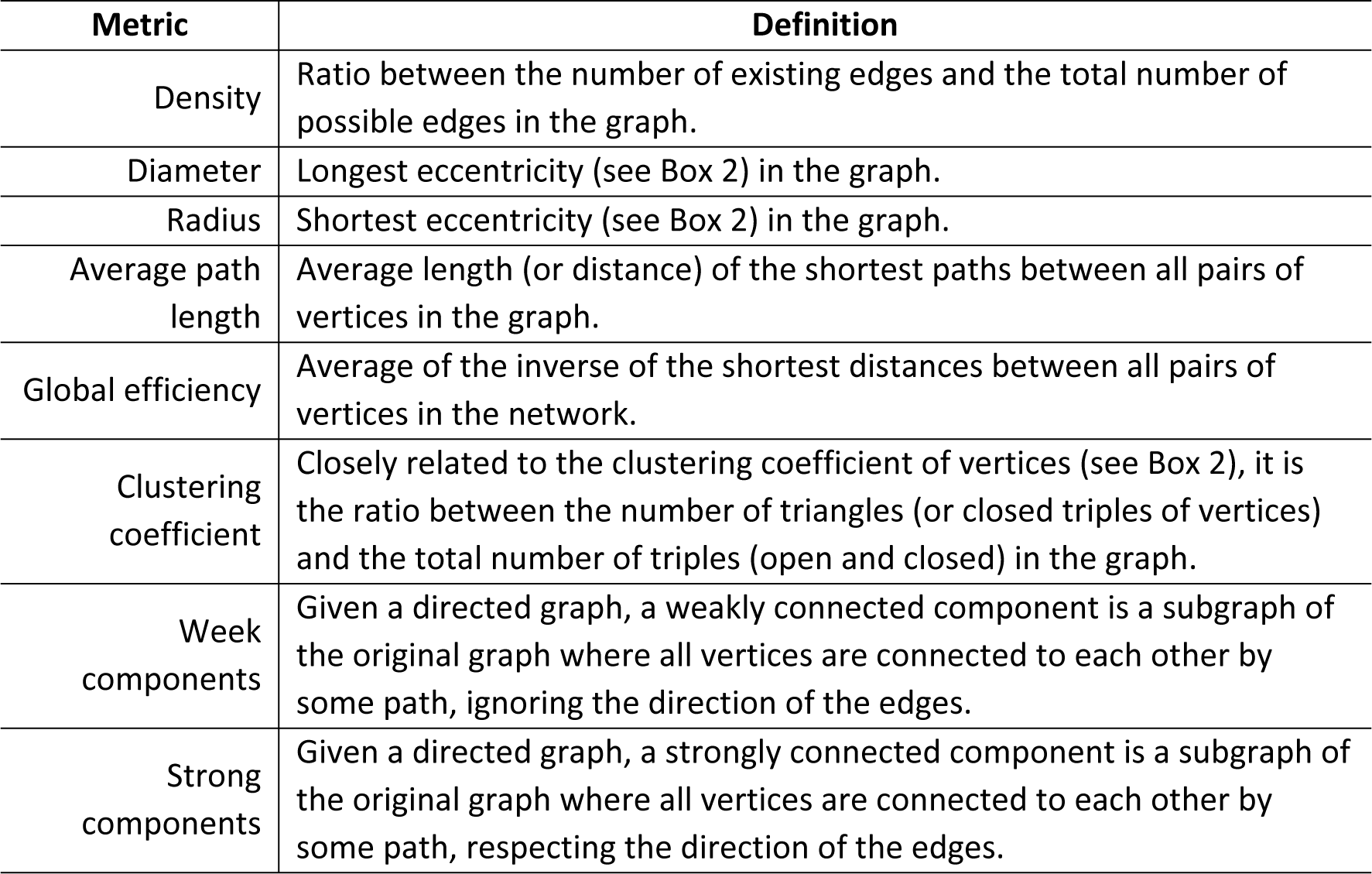

For the vertices, the following measures were calculated: referrals made by the physician; referrals received by the physician; follow-up consultations performed by the physician; degree-in; degree-out; clustering coefficient; local efficiency; closeness-in; closeness-out; betweenness; eccentricity; PageRank (Google); subgraph centrality; Kleinberg’s authority score; Kleinberg’s HUB score; diversity [16]. The calculation of weighted versions of the metrics was prioritized, giving the edges the weight Ew, as previously described. Unweighted versions were also calculated for some metrics for descriptive purposes or when the calculation of the weighted version was not applicable (Box 2).

##### Box 2. Network-level performance metrics

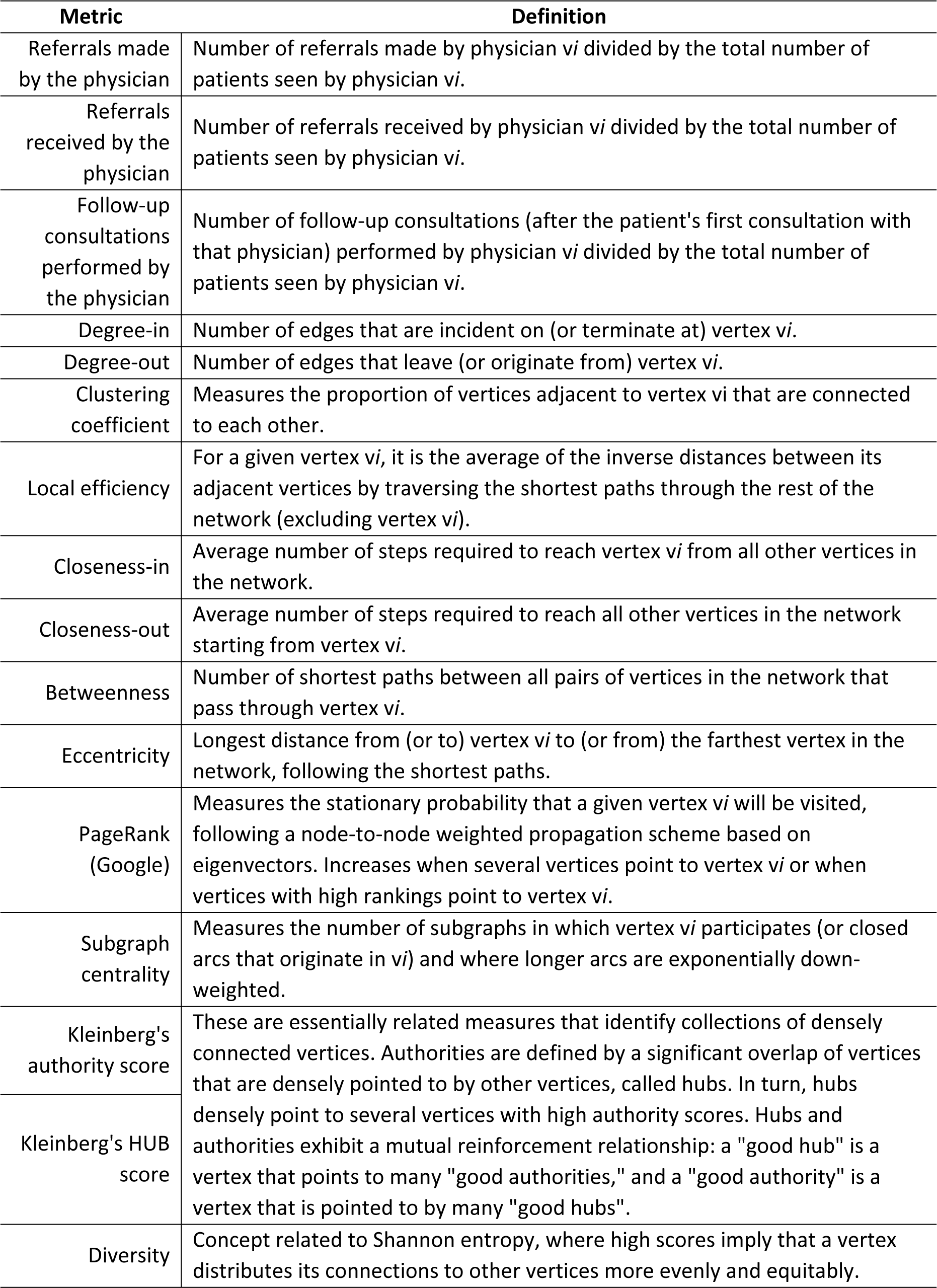

To ease comparisons between metrics with different dimensionalities, the results were expressed in terms of the number of standard deviations above or below the mean of the analyzed group.

Vertices that acted as articulation points (also called cut vertices) in the network were identified, defined as the vertices v*i* that, if removed, would increase the number of connected components in the graph or make a connected graph disconnected. Articulation points represent vulnerabilities for a connected network [16].

The vertex metrics (Box 2) were grouped into three dimensions (or constructs) according to the theoretical attributes they presumably reflect for the network and that are strategically significant from a managerial perspective. They are: a) “patient follow-up profile” dimension: aimed at characterizing whether the doctor practices a pattern of longitudinal follow-up or a more “episodic care” or “fragmented care” follow-up (i.e., where patients are seen on a sporadic or disconnected basis rather than consistently over an extended period of time). Represented by the candidate metrics degree-in, degree-out, referrals made, referrals received, and follow-up consultations; b) “relationship with authorities” dimension: aimed at characterizing the degree to which each doctor achieves high authority scores and/or contributes to raising the authority score of other doctors to whom they refer patients. Represented by Kleinberg’s authority and HUB scores; c) “centrality” dimension: aimed at positioning the doctor relative to the network graph, assuming that the more central the position of a doctor in the network, the greater their ability to access and disseminate knowledge and information, or in other words, control the flow of information and influence the patient’s care trajectory. Represented by the candidate metrics closeness-in, closeness-out, betweenness, eccentricity, PageRank, and subgraph centrality.

To characterize each physician according to the three profiles above, a cluster analysis was conducted using the non-hierarchical K-means technique, choosing the optimal number of clusters through visual inspection of graphs constructed by the methods of average silhouette width and within-clusters sum of squares. The physician clusters resulting from the network metrics were evaluated for their theoretical significance, and qualitative profiles (personas) were created for each dimension. For the “centrality” dimension: central, intermediary, or peripheral. For the “relationship with authorities” dimension: is authority, seeks authority, or balanced. For the “patient follow-up” dimension: strong prevalent, strong shared, medium shared, and weak shared.

The statistical association was evaluated between the centrality, relationship with authorities, and patient follow-up profiles, as well as the number of chronic comorbidities, emergency department visits, cost of exams (ordered by the analyzed physician), hospital admissions due to a medical condition, and total days of hospitalization due to a medical condition of the patients they attended. The association between categorical variables in contingency tables with dimensions greater than 2x2 was evaluated using the chi-square test and simple correspondence analysis with adjusted standardized residuals. Numerical variables were compared using the Kruskal-Wallis test, Mann-Whitney test, or t-test, as appropriate. Correlations between numerical variables were evaluated using the Pearson correlation coefficient (*r*).

The significance level was set at α = 0.05 (two-tailed). When necessary, the overall significance level was adjusted by means of Bonferroni correction for multiple comparisons.

Medical community detection was performed using the Infomap algorithm. This algorithm uses an information theoretic approach that is suitable for revealing community structures in weighted and directed networks. It uses the probability flow of random walks on a network as a proxy for information flows in the system and decompose the network into modules by compressing the probability flow. A group of nodes among which information flows quickly and easily (in our case, more dense referrals and counter-referrals) can be aggregated and described as a single well-connected module or community [16]. For this community detection procedure, edges and vertices were assigned weights Ew and Vw, previously described.

Due to the strong imbalance in the number of consultations among physicians, a definition was established for “low consultation productivity” when a physician had performed less than 20% of the consultations expected for his or her specialty, according to the criterion below:

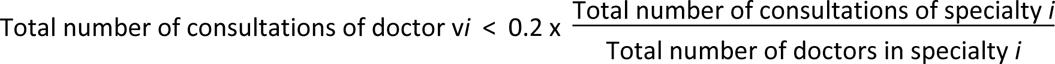

Doctors with low consultation productivity were included in the network-level SNA analysis but were excluded from the vertex-level performance analyses.

SNA and clustering analyses were conducted using the igraph and factoextra packages of R 4.2.0 language (R Core Team (2022). R: A language and environment for statistical computing. R Foundation for Statistical Computing, Vienna, Austria. URL https://www.R-project.org/) in the RStudio environment (Posit team (2022). RStudio: Integrated Development Environment for R. Posit Software, PBC, Boston, MA. URL http://www.posit.co/). Other analyses, including data handling and pre-processing, were performed using Stata/SE 11.2 software (StataCorp LP, College Station, TX, USA).

The research was conducted following the principles of Brazilian ethical resolutions, particularly Resolution No. 466/12 of the National Health Council and its complementary resolutions. The project received approval from a research ethics committee endorsed by the National Commission for Ethics in Research (CONEP) (submission identifier No. 68241023.8.0000.5128. Collegiate decision No. 6.019.051).

## Results

### Network-level measures

During the study period, 666,263 individuals had at least one office visit, totaling 3,863,222 visits with one or more of the 4,554 physicians accredited by the PHIP. Only 15 physicians did not receive referrals or referred patients to other colleagues during the period and were excluded from SNA analyses. The results of the network-level measures are shown in Table 1.

**Table 1.** Results of the network-level measures.

|  |  |
| --- | --- |
| Number of vertices | 4,539 |
| Number of edges | 1,160,346 |
| Weak components | 1 |
| Strong components | 38 |
| Clustering coefficient | 0.255 |
| Density | 5.63% |
| Diameter | 5 (unweighed); 34.417 (weighed) |
| Radius | 3 (ignoring vertices with degree-out equal to zero) |
| Average path length | 2.048 (unweighed); 3.346 (weighed) |
| Global efficiency | 0.507 (unweighed); 0.339 (weighed) |
| Number of articulation point | 27 |
| Number of medical communities | 15 (modularity = 0.149) |

### Physicians with low consultation productivity

A total of 577 physicians (12.67%) were classified as having low consultation productivity (see Method). These physicians were responsible for only 18,058 referrals made (1.08%) and 17,961 referrals received (1.07%). The mean age of this group was 54.76 years-old (95% CI = 53.74 - 55.79), not statistically different from that of physicians above this consultation threshold (55.51 years-old, 95% CI = 55.18 - 55.84; t-test = 1.545, p = 0.123). Physicians with low consultation productivity had lower mean satisfaction scores than the rest [9.57 (95% CI = 9.51 - 9.64) vs. 9.65 (95% CI = 9.64 - 9.67), t-test = 3.92, p = 0.001].

The distribution of physicians according to consultation productivity and medical specialty is shown in S1 Table. The specialties most strongly associated with low consultation productivity were anesthesiology, general surgery, internal medicine, endoscopy, and family and community medicine.

### Vertex-level measures

All subsequent analysis of vertex-level metrics was conducted by excluding these 577 physicians with low consultation productivity, leaving 3,977 professionals. Distribution of vertex metric results showed strong variations among physicians, suggesting that those metrics may in fact be capturing different roles for each doctor in the network (Table 2).

**Table 2.** Results of vertex-level measures.

| Measure | Number of standard deviations above or below the mean |  |  |  |  |
| --- | --- | --- | --- | --- | --- |
|  | Minimum | 25<sup>th</sup><br>percentile | Median | 75<sup>th</sup><br>percentile | Maximum |
| Referrals made by the physician | -2.04 | -0.56 | -0.15 | 0.35 | 17.23 |
| Referrals received by the physician | -2.65 | -0.60 | -0.10 | 0.42 | 16.08 |
| Follow-up consultations performed by the physician | -1.37 | -0.61 | -0.23 | 0.29 | 10.45 |
| Degree-in (unweighted) | -1.35 | -0.76 | -0.27 | 0.53 | 5.35 |
| Degree-in (weighted) | -1.23 | -0.70 | -0.29 | 0.44 | 8.14 |
| Degree-out (unweighted) | -1.32 | -0.76 | -0.27 | 0.53 | 5.40 |
| Degree-out (weighted) | -2.04 | -0.56 | -0.15 | 0.35 | 17.23 |
| Clustering coefficient (unweighted) | -3.03 | -0.58 | -0.08 | 0.49 | 7.79 |
| Clustering coefficient (weighted) | -2.82 | -0.61 | -0.12 | 0.44 | 7.17 |
| Local efficiency (unweighted) | -2.98 | -0.53 | -0.15 | 0.33 | 11.79 |
| Local efficiency (weighted) | -1.08 | -0.50 | -0.37 | -0.09 | 9.33 |
| Closeness-in (unweighted) | -6.88 | -0.53 | 0.03 | 0.62 | 3.70 |
| Closeness-in (weighted) | -5.30 | -0.69 | -0.02 | 0.66 | 3.78 |
| Closeness-out (unweighted) | -4.89 | -0.57 | 0.02 | 0.63 | 3.82 |
| Closeness-out (weighted) | -2.79 | -0.69 | -0.01 | 0.65 | 4.30 |
| Betweenness (unweighted) | -0.62 | -0.54 | -0.36 | 0.13 | 13.12 |
| Betweenness (weighted) | -0.41 | -0.38 | -0.28 | -0.03 | 22.56 |
| Eccentricity | -0.69 | -0.69 | -0.69 | 1.45 | 1.45 |
| PageRank (Google) | -1.20 | -0.70 | -0.30 | 0.41 | 7.31 |
| Subgraph centrality | -0.75 | -0.65 | -0.40 | 0.30 | 7.32 |
| Kleinberg's authority score | -0.09 | -0.07 | -0.05 | -0.02 | 45.47 |
| Kleinberg's HUB score | -1.13 | -0.50 | -0.24 | 0.16 | 16.49 |
| Diversity | -10.14 | -0.28 | 0.26 | 0.61 | 1.28 |

### Centrality dimension measures

There was a strong linear correlation among several candidate metrics of the centrality dimension. This led to considering weighted closeness-out, weighted closeness-in, eccentricity, and PageRank (Google) metrics in the clustering analysis (S2 Table). Subgraph centrality, weighted betweenness, and unweighted betweenness did not participate in the clustering but their distribution in the emerging clusters will also be shown, endorsing the stated correlation. The cluster analysis suggested the existence of three clusters, which were named as “central”, “intermediate”, and “peripheral” summary profiles (Table 3).

**Table 3.** Cluster analysis results for centrality dimension metrics, in number of standard deviations above or he mean.

| Cluster | N | Weighted closeness-out* | Weighted closeness-in* | Eccentricity* | PageRank (Google)* | Subgraph centrality** | Unweighted between-ness** | Weighted between-ness** | Summary profile |
| --- | --- | --- | --- | --- | --- | --- | --- | --- | --- |
| 1 | 1,768 | -0.1 | -0.2 | -0.7 | -0.3 | -0.3 | -0.3 | -0.1 | Intermediate |
| 2 | 1,258 | +0.6 | -0.6 | +1.5 | -0.7 | -0.6 | -0.5 | 0.0 | Peripheral |
| 3 | 951 | -0.7 | +1.1 | -0.6 | +1.4 | +1.3 | +1.2 | +0.1 | Central |

\*Included in clustering analysis.
\*\*Not included in clustering analysis.

The distribution of physicians according to their centrality profile and specialty is shown in Table 4. Cardiology, dermatology, endocrinology, ophthalmology, orthopedics, otolaryngology, pulmonology, psychiatry, and urology were strongly associated with the central profile. Surgical specialties predominated in the peripheral position, as well as clinical specialties such as nephrology, infectious diseases, internal medicine, and pediatrics.

**Table 4.**
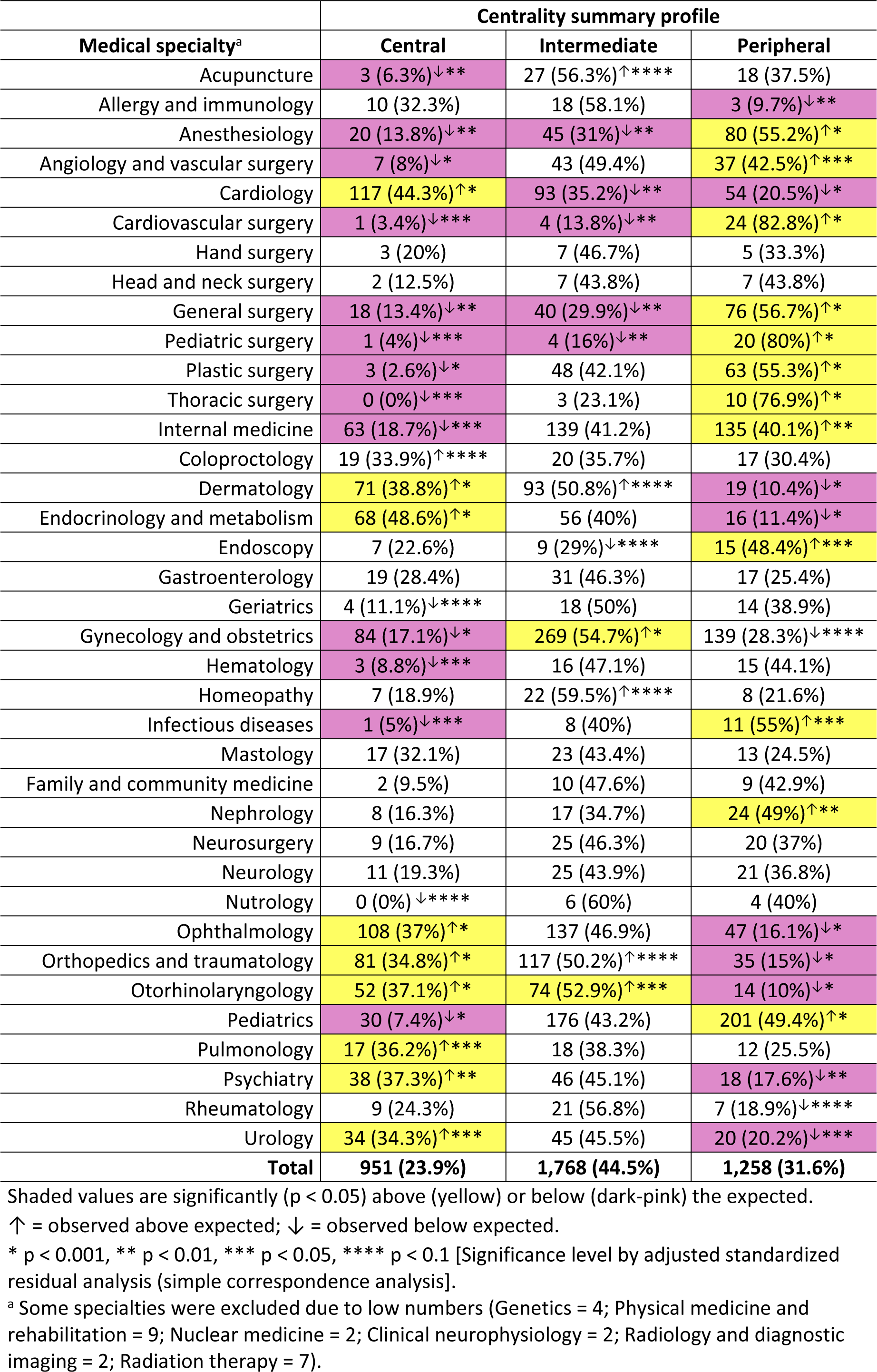
Distribution of physicians according to their centrality profile and specialty.

### Relationship with authorities dimension measures

The variables included in the clustering were Kleinberg’s authority and HUB scores. The cluster analysis suggested the existence of four clusters which were summarized into three, named as “is authority”, “seeks authorities”, and “balanced” profiles (Table 5).

**Table 5.** Cluster analysis results for the authority dimension metrics, in number of standard deviations above or below the mean.

| Cluster | N | Kleinberg's authority score | Kleinberg's HUB score | Summary profile |
| --- | --- | --- | --- | --- |
| 1 | 233 | -0.04 | +2.1 | Seeks authorities |
| 2 | 883 | +0.15 | +0.5 | Is authority |
| 3 | 2,828 | -0.04 | -0.4 | Balanced |
| 4 | 33 | +0.10 | +6.9 | Seeks authorities |

The distribution of physicians according to their authority profile and specialty is shown in Table 6. There was a higher presence of physicians with an “is authority” profile in specialties such as cardiology, endocrinology, gastroenterology, geriatrics, homeopathy, nephrology, neurology, nutrology, pulmonology, psychiatry, and rheumatology.

**Table 6.**
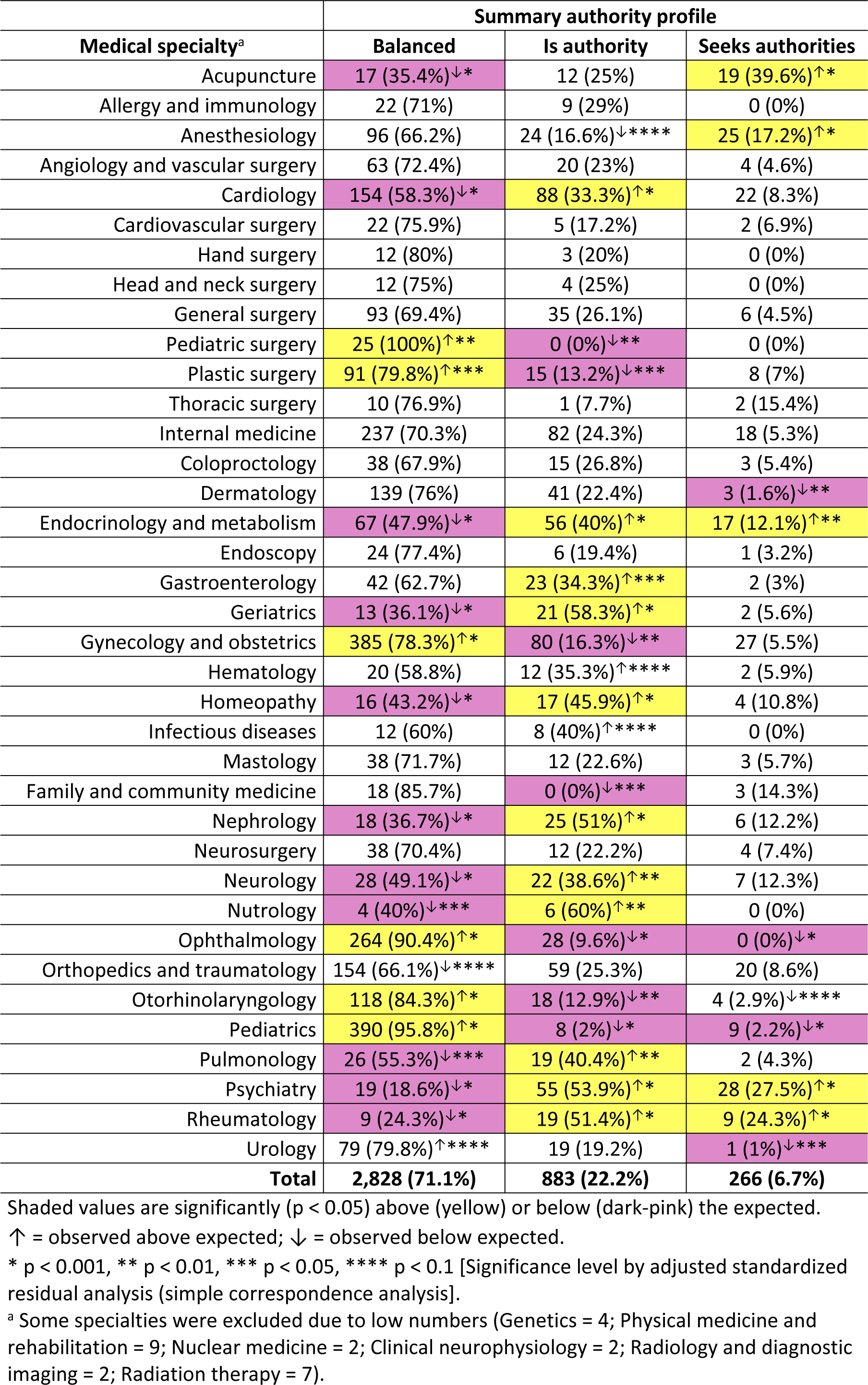
Distribution of physicians according to their authority profile and specialty.

### Patient follow-up dimension measures

There was a strong linear correlation among several candidate metrics in the patient follow-up dimension. This led to including number of referrals made (per patient seen), number of referrals received (per patient seen), and number of follow-up appointments (per patient seen) in the clustering (S2 Table). Degree-in and degree-out, both weighted and unweighted, did not participate in the clustering, but their distribution in the clusters found will also be reported, corroborating the quoted correlation. The cluster analysis suggested the existence of five clusters, which were summarized into four, named as “strong, shared”, “medium, shared”, “weak, shared”, and “strong, prevalent” profiles (Table 7).

**Table 7.** Cluster analysis results for patient follow-up dimension metrics, in number of standard deviations above or below the mean.

| Cluster | N | Referrals made* | Referrals received* | Follow-up appointments* | Unweighted degree-in** | Weighted degree-in** | Unweighted degree-out** | Weighted degree-out** | Summary profile |
| --- | --- | --- | --- | --- | --- | --- | --- | --- | --- |
| 1 | 64 | +4.7 | +4.5 | +4.4 | +0.1 | +0.2 | +0.1 | +4.7 | Strong, shared |
| 2 | 1,502 | -0.6 | -0.6 | -0.6 | -0.0 | -0.1 | -0.0 | -0.6 | Weak, shared |
| 3 | 293 | -0.9 | -1.0 | +1.2 | -0.8 | -0.5 | -0.8 | -0.9 | Strong, prevalent |
| 4 | 455 | +1.3 | +1.3 | +1.1 | +0.2 | +0.3 | +0.2 | +1.3 | Strong, shared |
| 5 | 1,663 | +0.2 | +0.2 | -0.1 | +0.1 | +0.1 | +0.1 | +0.2 | Medium, shared |

\*Included in clustering analysis.
\*\*Not included in clustering analysis.

The distribution of physicians according to their patient follow-up profile and specialty is shown in Table 8. The specialties significantly associated with strong and shared patient follow-up were acupuncture, internal medicine, endocrinology, geriatrics, hematology, homeopathy, nephrology, neurology, nutrology, psychiatry, and rheumatology. Only pediatrics was significantly associated with strong and prevalent patient follow-up. There was a significant association between weak and shared follow-up for internal medicine and family medicine, among other specialties.

**Table 8.**
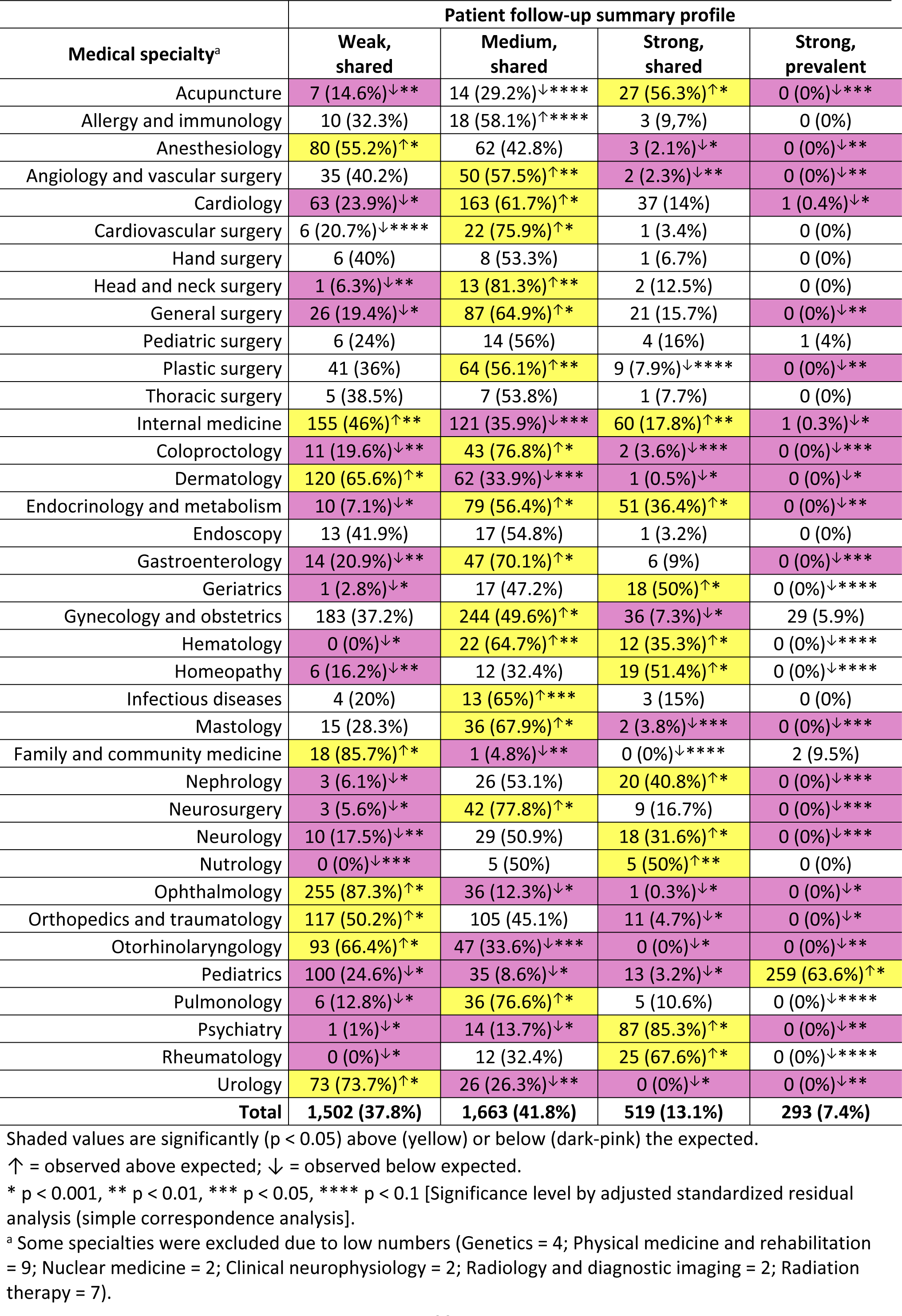
Distribution of physicians according to their patient follow-up profile and specialty.

### Associations between physician summary profiles

Each of the created profiles (i.e., centrality, patient follow-up, and relationship with authorities) exhibited a strong and statistically significant pairwise relationship with the other profiles (chi-square test, p < 0.001 for all pairwise comparisons).

Physicians in a central position were more likely to have a strong and shared patient follow-up profile, while physicians in a peripheral position were associated with a strong and prevalent follow-up profile. Physicians in an intermediate position exhibited a non-statistically significant tendency to weak and shared patient follow-up (Table 9).

**Table 9.** Association between patient follow-up and centrality profiles of doctors.

| Centrality summary profile | Patient follow-up summary profile |  |  |  | Total |
| --- | --- | --- | --- | --- | --- |
|  | Weak, shared | Medium, shared | Strong, shared | Strong, prevalent |  |
| **Central** | 346 | 420 | 162 | 23 | 951 (23.9%) |
| % of the line | 36.4% | 44.2%<sup>↑****</sup> | 17.0%<sup>↑*</sup> | 2.4%<sup>↓*</sup> |  |
| % of the column | 23.0% | 25.3%<sup>↑****</sup> | 31.2%<sup>↑*</sup> | 7.8%<sup>↓*</sup> |  |
| **Intermediate** | 695 | 723 | 215 | 135 | 1.768 (44.5%) |
| % of the line | 39.3%<sup>↑****</sup> | 40.9% | 12.2% | 7.6% |  |
| % of the column | 46.3%<sup>↑****</sup> | 43.5% | 41.4% | 46.1% |  |
| **Peripheral** | 461 | 520 | 142 | 135 | 1.258 (31.6%) |
| % of the line | 36.6% | 41.3% | 11.3%<sup>↓***</sup> | 10.7%<sup>↑*</sup> |  |
| % of the column | 30.7% | 31.3% | 27.4%<sup>↓***</sup> | 46.1%<sup>↑*</sup> |  |
| **Total** | **1,502 (37.8%)** | **1,663 (41.8%)** | **519 (13.1%)** | **293 (7.4%)** | **3,977** |

Shaded values are significantly ( $p < 0.05$ ) above (yellow) or below (dark-pink) the expected.
↑ = observed above expected; ↓ = observed below expected.
\* $p < 0.001$ , \*\*\* $p < 0.05$ , \*\*\*\* $p < 0.1$ [Significance level by adjusted standardized residual analysis (simple correspondence analysis)].

Authorities were more likely to be in central positions within the network, while physicians seeking authorities were more often located in intermediate positions in the network, next to them. Physicians located in the periphery more frequently had a balanced relationship with authorities (Table 10).

**Table 10.** Association between authority and centrality profiles of doctors.

|  | Relationship with authorities summary profile |  |  |  |
| --- | --- | --- | --- | --- |
| Centrality summary profile | Balanced | Is authority | Seeks authorities | Total |
| **Central** | 572 | 345 | 34 | 951 (23.9%) |
| % of the line | 60.1%↓* | 36.3%↑* | 3.6%↓* |  |
| % of the column | 20.2%↓* | 39.1%↑* | 12.8%↓* |  |
| **Intermediate** | 1,283 | 351 | 134 | 1,768 (44.5%) |
| % of the line | 72.6%↑**** | 19.9%↓** | 7.6%↑*** |  |
| % of the column | 45.4%↑**** | 39.8%↓** | 50.4%↑*** |  |
| **Peripheral** | 973 | 187 | 98 | 1,258 (31.6%) |
| % of the line | 77.3%↑* | 14.9%↓* | 7.8%↑**** |  |
| % of the column | 34.4%↑* | 21.2%↓* | 36.8%↑**** |  |
| **Total** | **2,828 (71.1%)** | **883 (22.2%)** | **266 (6.7%)** | **3,977** |

Shaded values are significantly ( $p < 0.05$ ) above (yellow) or below (dark-pink) the expected.
↑ = observed above expected; ↓ = observed below expected.
\* $p < 0.001$ , \*\* $p < 0.01$ , \*\*\* $p < 0.05$ , \*\*\*\* $p < 0.1$ [Significance level by adjusted standardized residual analysis (simple correspondence analysis)].

Authorities more frequently exhibited strong and medium shared patient follow-up profiles. Physicians with a tendency to seek authorities were more likely to have a strong and shared patient follow-up profile. Lastly, a weak follow-up profile was characteristic of physicians not classified as authorities or seeking authorities (Table 11).

**Table 11.** Association between authority and patient follow-up profiles of doctors.

|  | Relationship with authorities summary profile |  |  |  |
| --- | --- | --- | --- | --- |
| Patient follow-up summary profile | Balanced | Is authority | Seeks authorities | Total |
| **Weak, shared** | 1.310 | 153 | 39 | 1,502 (37.8%) |
| % of the line | 87.2%<sup>↑*</sup> | 10.2%<sup>↓*</sup> | 2.6%<sup>↓*</sup> |  |
| % of the column | 46.3%<sup>↑*</sup> | 17.3%<sup>↓*</sup> | 14.7%<sup>↓*</sup> |  |
| **Medium, shared** | 1.109 | 428 | 126 | 1,663 (41.8%) |
| % of the line | 66.7%<sup>↓*</sup> | 25.7%<sup>↑*</sup> | 7.6%<sup>↑****</sup> |  |
| % of the column | 39.2%<sup>↓*</sup> | 48.5%<sup>↑*</sup> | 47.4%<sup>↑****</sup> |  |
| **Strong, shared** | 130 | 295 | 94 | 519 (13.1%) |
| % of the line | 25.0%<sup>↓*</sup> | 56.8%<sup>↑*</sup> | 18.1%<sup>↑*</sup> |  |
| % of the column | 4.6%<sup>↓*</sup> | 33.4%<sup>↑*</sup> | 35.3%<sup>↑*</sup> |  |
| **Strong, prevalent** | 279 | 7 | 7 | 293 (7.4%) |
| % of the line | 95.2%<sup>↑*</sup> | 2.4%<sup>↓*</sup> | 2.4%<sup>↓**</sup> |  |
| % of the column | 9.9%<sup>↑*</sup> | 0.8%<sup>↓*</sup> | 2.6%<sup>↓**</sup> |  |
| **Total** | **2,828 (71.1%)** | **883 (22.2%)** | **266 (6.7%)** | **3,977** |

Shaded values are significantly ( $p < 0.05$ ) above (yellow) or below (dark-pink) the expected.
↑ = observed above expected; ↓ = observed below expected.
\* $p < 0.001$ , \*\* $p < 0.01$ , \*\*\*\* $p < 0.1$ [Significance level by adjusted standardized residual analysis (simple correspondence analysis)].

### Association of physician profiles with patient clinical characteristics and outcomes

Patients with a higher number of chronic comorbidities showed a progressive increase in the risk of hospitalization due to a medical condition (*r* = 0.545, p < 0.001), hospitalization for primary care-sensitive conditions (*r* = 0.402, p < 0.001), and days of hospitalization due to a medical condition (*r* = 0.547, p < 0.001), but not emergency department visits (*r* = 0.012, p = 464). The correlation between the number of comorbidities and the cost of exams was weak but statistically significant (*r* = 0.221, p < 0.001).

The three physician profiles also showed statistically significant associations with the clinical characteristics and outcomes of the patients they treated (Table 12).

**Table 12.** Associations between physician profiles and the clinical characteristics and outcomes of the patients they treated.

| Physician summary profiles | Number of chronic comorbidities (per patient)* | Emergency department visits (per patient)* | Number of hospital admissions (per 100 patients)* | Total days of hospitalization (per 100 patients)* | Total cost with ordered exams (US\$)*<sup>a</sup> |
| --- | --- | --- | --- | --- | --- |
| **Centrality<sup>b</sup>** |  |  |  |  |  |
| Central | 1.92<br>(1.41 - 2.47) | 1.55<br>(1.35 - 1.80) | 4.89<br>(3.32 - 7.55) | 40.50<br>(26.89 - 66.76) | 35.7<br>(16.5 - 56.5) |
| Intermediate | 1.77<br>(1.25 - 2.37) | 1.64<br>(1.39 - 1.95) | 4.93<br>(3.00 - 8.00) | 38.17<br>(20.92 - 68.72) | 37.0<br>(15.5 - 64.8) |
| Peripheral | 1.89<br>(1.17 - 2.67) | 1.67<br>(1.38 - 2.08) | 5.88<br>(3.36 - 10.83) | 44.61<br>(21.98 - 90.91) | 34.5<br>(14.8 - 65.6) |
| p-value (Kruskal-Wallis) | < 0.001 | < 0.001 | < 0.001 | < 0.001 | 0.142 |
| **Relationship with authorities<sup>b</sup>** |  |  |  |  |  |
| Balanced | 1.69<br>(1.15 - 2.29) | 1.64<br>(1.38 - 2.00) | 4.88<br>(3.62 - 8.14) | 37.40<br>(21.50 - 66.59) | 32.8<br>(15.0 - 59.5) |
| Is authority | 2.24<br>(1.71 - 2.91) | 1.57<br>(1.35 - 1.80) | 6.00<br>(3.60 - 10.22) | 51.34<br>(27.60 - 91.45) | 44.4<br>(20.8 - 69.6) |
| Seeks authorities | 2.25<br>(1.67 - 2.91) | 1.62<br>(1.41 - 1.94) | 6.13<br>(3.81 - 9.38) | 46.75<br>(25.33 - 84.14) | 42.1<br>(8.4 - 71.8) |
| p-value (Kruskal-Wallis) | < 0.001 | < 0.001 | < 0.001 | < 0.001 | < 0.001 |
| **Patient follow-up<sup>c</sup>** |  |  |  |  |  |
| Weak, shared | 1.60<br>(1.23 - 2.02) | 1.59<br>(1.36 - 1.89) | 4.13<br>(2.83 - 6.15) | 33.62<br>(20.09 - 52.32) | 26.4<br>(11.3 - 49.4) |
| Medium, shared | 2.17<br>(1.61 - 2.81) | 1.61<br>(1.37 - 1.91) | 5.76<br>(3.45 - 9.92) | 46.57<br>(25.24 - 87.43) | 46.2<br>(27.0 - 71.3) |
| Strong, shared | 2.49<br>(1.84 - 3.30) | 1.60<br>(1.36 - 1.93) | 7.19<br>(4.25 - 12.83) | 62.07<br>(31.30 - 116.41) | 48.4<br>(12.6 - 90.7) |
| Strong, prevalent | 0.38<br>(0.28 - 0.57) | 1.98<br>(1.64 - 2.37) | 6.26<br>(4.17 - 9.01) | 34.19<br>(18.93 - 56.47) | 14.5<br>(10.0 - 25.7) |
| p-value (Kruskal-Wallis) | < 0.001 | < 0.001 | < 0.001 | < 0.001 | < 0.001 |

\* Values are medians (25<sup>th</sup> - 75<sup>th</sup> percentiles).
<sup>a</sup> The values were calculated in Brazilian local currency and converted into US dollars, based on the exchange rate in April 2023.
<sup>b</sup> Values shaded in yellow are statistically different (significance level < 0.05/2 = 0.025) from dark-pink-shaded values, and vice-versa.
<sup>c</sup> Values shaded in orange are statistically different (significance level < 0.05/3 = 0.017) from all other three categories. Values shaded in blue are statistically different (significance level < 0.05/3 = 0.017) only from orange-shaded values.
All pairwise comparisons by Mann-Whitney test.

As to the centrality profile, physicians in central positions attended patients with the highest number of chronic comorbidities, but these patients were less likely to visit emergency services. Physicians in intermediate positions attended patients with fewer chronic comorbidities and who spent less days at hospital. Patients attended by physicians in peripheral positions had the highest incidence of hospitalization and hospital stay (Table 12). There were no statistically significant differences in the cost of ambulatory exams per patient ordered by the doctor.

Physicians with a tendency to seek authorities and physicians who are authorities attended patients with the highest number of comorbidities, but patients assisted by authorities visited emergency services the least. Physicians with a balanced relationship with authorities attended patients with significantly fewer comorbidities, less hospitalizations and hospital stay, and had the lowest costs of exams per patient (Table 12).

Amongst physicians with shared follow-up standards, the presence of chronic comorbidities, incidence of hospitalization and total days of hospital stay increased as the intensity of patient follow-up increased, but there was no relationship with emergency service visits. Physicians with weak follow-up standards generated lower costs with tests and therapies per patient. Patients cared for by physicians with strong and prevalent follow-up profile showed the lowest number of comorbidities and costs with tests and therapies, but the highest utilization of emergency services (Table 12).

### Articulation points

Twenty-seven physicians were identified as behaving as articulation points in the network, with the following characteristics: four cardiologists, three each of endocrinologists, ophthalmologists, and pediatricians, with the remaining 14 being in smaller numbers from other specialties. Ten doctors were in a central position in the network and 10 were in an intermediate position, 13 were authorities, and 14 had a medium and shared patient follow-up profile.

### Medical communities

Overall, 15 medical communities were identified when considering the 4,539 physicians who received or referred at least one patient to another colleague during the period (Table 1). Four of these communities were small clusters of physicians classified as having low consultation productivity (S1 Table). After excluding these, 11 communities were detected (Table 13).

**Table 13.** Number of physicians per identified community, according to their medical specialty.

| Medical specialty<sup>a</sup> | Medical community |  |  |  |  |  |  |  |  |  | Total |
| --- | --- | --- | --- | --- | --- | --- | --- | --- | --- | --- | --- |
|  | A | B | C | D | E | F | G | H | I | J |  |
| Acupuncture | 43 | 3 | - | 1 | 1 | - | - | - | - | - | 48 |
| Allergy and immunology | 25 | 1 | 4 | - | 1 | - | - | - | - | - | 31 |
| Anesthesiology | 141 | 2 | 1 | 1 | - | - | - | - | - | - | 145 |
| Angiology and vascular surgery | 82 | 3 | - | 2 | - | - | - | - | - | - | 87 |
| Cardiology | 245 | 11 | 2 | 4 | 1 | - | 1 | - | - | - | 264 |
| Cardiovascular surgery | 28 | 1 | - | - | - | - | - | - | - | - | 29 |
| Hand surgery | 15 | - | - | - | - | - | - | - | - | - | 15 |
| Head and neck surgery | 16 | - | - | - | - | - | - | - | - | - | 16 |
| General surgery | 126 | 6 | - | 2 | - | - | - | - | - | - | 134 |
| Pediatric surgery | - | - | 22 | 1 | - | 2 | - | - | - | - | 25 |
| Plastic surgery | 110 | 3 | - | - | 1 | - | - | - | - | - | 114 |
| Thoracic surgery | 12 | - | 1 | - | - | - | - | - | - | - | 13 |
| Internal medicine | 299 | 29 | - | 3 | 6 | - | - | - | - | - | 337 |
| Coloproctology | 55 | 1 | - | - | - | - | - | - | - | - | 56 |
| Dermatology | 174 | 6 | 1 | 1 | 1 | - | - | - | - | - | 183 |
| Endocrinology and metabolism | 131 | 5 | 1 | 2 | 1 | - | - | - | - | - | 140 |
| Endoscopy | 25 | 3 | 1 | - | 1 | - | 1 | - | - | - | 31 |
| Gastroenterology | 64 | 3 | - | - | - | - | - | - | - | - | 67 |
| Medical genetics | 1 | - | 3 | - | - | - | - | - | - | - | 4 |
| Geriatrics | 34 | 2 | - | - | - | - | - | - | - | - | 36 |
| Gynecology and obstetrics | 451 | 24 | - | 7 | 7 | - | 3 | - | - | - | 492 |
| Hematology | 33 | - | 1 | - | - | - | - | - | - | - | 34 |
| Homeopathy | 32 | 2 | 2 | - | 1 | - | - | - | - | - | 37 |
| Infectious diseases | 20 | - | - | - | - | - | - | - | - | - | 20 |
| Mastology | 52 | 1 | - | - | - | - | - | - | - | - | 53 |
| Family and community medicine | 18 | 2 | - | - | - | - | 1 | - | - | - | 21 |
| Physical medicine and rehab | 7 | - | - | - | - | - | - | - | - | - | 7 |
| Nephrology | 46 | 1 | 2 | - | - | - | - | - | - | - | 49 |
| Neurosurgery | 49 | 1 | 4 | - | - | - | - | - | - | - | 54 |
| Neurology | 52 | 3 | - | 1 | 1 | - | - | - | - | - | 57 |
| Nutrology | 10 | - | - | - | - | - | - | - | - | - | 10 |
| Ophthalmology | 267 | 9 | 9 | 4 | 1 | - | 1 | - | - | 1 | 292 |
| Orthopedics and traumatology | 211 | 11 | 6 | 1 | 3 | - | 1 | - | - | - | 233 |
| Otorhinolaryngology | 124 | 6 | 5 | 2 | 1 | - | 2 | - | - | - | 140 |
| Pediatrics | 10 | 1 | 347 | 3 | 6 | 29 | 2 | 6 | 2 | 1 | 407 |
| Pulmonology | 42 | 4 | - | 1 | - | - | - | - | - | - | 47 |
| Psychiatry | 100 | 1 | 1 | - | - | - | - | - | - | - | 102 |
| Radiation therapy | 7 | - | - | - | - | - | - | - | - | - | 7 |
| Rheumatology | 37 | - | - | - | - | - | - | - | - | - | 37 |
| Urology | 90 | 8 | - | - | 1 | - | - | - | - | - | 99 |
| **Total** | **3,288** | **153** | **413** | **36** | **34** | **31** | **12** | **6** | **2** | **2** | **3,977** |

<sup>a</sup> Some specialties were excluded due to low numbers (Nuclear medicine = 2; Clinical neurophysiology = 2; Radiology and diagnostic imaging = 2).

Box 3 shows a clear territorial and medical specialty segregation within the emerging communities. It is noteworthy that no community was detected with a primary seat in Contagem, the second most populous municipality in the area covered by our PHIP.

##### Box 3. Characteristics of identified communities of physicians

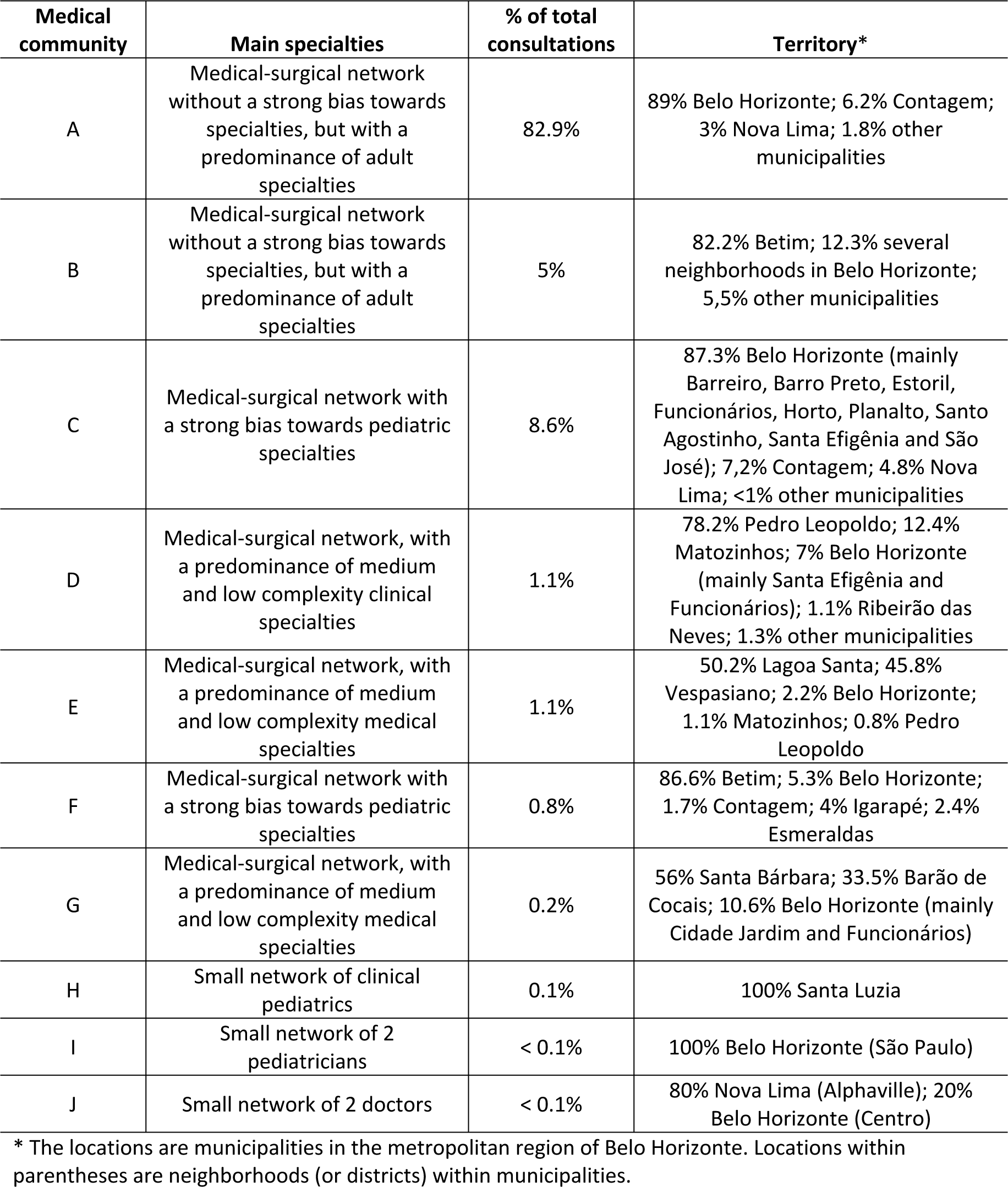

## Discussion

This study aimed to characterize the structure of the Amb-HCN of a PHIP located in Belo Horizonte, Southern Brazil, by applying objective measures from graph theory to the patient referral and counter-referral flows among the attending physicians. Specifically, it was possible to identify medical specialties with different standards of centrality in the network, their relationship with authorities, and patient follow-up. The study demonstrated the relationship between these different profiles and their association with patient clinical characteristics and outcomes. Importantly, it characterized the most frequent profiles for each medical specialty and the significant dispersion of physician behaviors within each specialty.

Understanding the structure and organic functioning of an Amb-HCN involves understanding the roles and responsibilities of physicians during and after patient care in their offices. The presence of shared patients between two or more physicians reveals relationships that are established explicitly (i.e., established in contracts and formally monitored through performance indicators and value delivery), informally (i.e., established naturally and spontaneously, due to physician’s and patient’s preferences, sociodemographic characteristics, terrain, etc.), or even by chance, but represent a valuable source of information for the study of care networks [4, 9]. Physicians establish patient referral bonds more frequently with other physicians of the same sex and age group when working in the same institution or geographically close, when completed their degree or residency training at the same educational institution, and when treating patients with similar clinical complexity, among other factors [17]. In any case, managing the functioning of an Amb-HCN presupposes the identification and measurement of these roles and responsibilities of the actors involved in direct patient care. Studies support the use of graph theory and SNA metrics to express the structure of healthcare networks, explain their care outcomes, study their changes over time, and observe how they react to dynamic influences of central governance policies [9, 18, 19].

Studies suggest that various structural properties of healthcare networks may be associated with quality and safety of care [9, 20], although there remains a vast field of research to be explored. The dimensions used in this study to characterize the Amb-HCN were defined based on attributes considered strategically important for health policy makers [9, 20, 21]. The proposed analysis in this study aimed to identify the professionals who occupy prominent positions in the network, either due to their relationships with their peers, their connections with influential physicians, or because the topology of the network would change substantially without them. Thus, the following components were identified as key factors for identifying prominent professionals: patient referrals received from peers, relative importance in the network, and patient return behavior. Similar criteria were followed in other studies [22].

Regarding the centrality dimension, it is assumed that doctors occupying more central positions in the graph have access to the most intense flow of information from colleagues who preceded them in patient care, and their own conduct can significantly influence the conduct of colleagues who will succeed them in patient care [7, 18, 20, 21]. Measures of centrality quantify the ability of a vertex to send, receive, or interrupt the flow of information [9]. Therefore, these doctors have a significant influence on the care trajectory of patients who seek them, even without conscious awareness or formally assuming this role [20, 21]. Another possible interpretation for the central role of some specialties is that they reflect the most prevalent nosology of the patient portfolio, which requires critical analysis by the health manager in the face of the central position of specialties such as cardiology, endocrinology, pulmonology, orthopedics, and psychiatry. Of particular interest is the central position occupied by urology, which may reflect the prevalent cultural practice in our setting of this professional assuming the health care of men in many situations. It may be argued that having many specialist doctors occupying the center of the network (as shown in Table 4) would not be the ideal structure for an Amb-HCN from the patient’s point of view, assuming that this position should be occupied by generalist and primary care physicians with the ability to coordinate patient care [10, 23]. Indeed, authors have proposed calculating the ratio between the centrality of primary care physicians and that of specialist physicians in the network [4, 23]. However, some studies have failed to demonstrate that Amb-HCN where primary care physicians were more central led to better health outcomes [23]. A study conducted with data from a private healthcare organization in Brazil found very similar centrality profiles to those of this study, with the most prevalent medical specialties being cardiology, endocrinology, dermatology, hematology, nephrology, orthopedics, and otorhinolaryngology [24]. Another analysis conducted in Amb-HCN in the German public health system, where patients can seek care directly from specialists without needing to go through a primary care physician, also observed a notable dispersion of specialties involved in the care of patients with chronic diseases: 72% of the networks involved at least 10 distinct specialties, and the physicians with greater centrality in the networks were more often specialists (e.g., otorhinolaryngology, ophthalmology, etc.) [10]. On the other hand, peripheral positions in our Amb-HCN are predominantly occupied by surgical specialties, which can be readily explained by the nature of these specialties. However, the peripheral position of internal medicine, nephrology, and pediatrics is remarkable. In the case of the latter two, it could be explained by their strong profile of longitudinal patient follow-up (Table 8), which would lead them to assume a large part of patient care and have few connections with other colleagues. It is necessary to understand whether the unexpected strong peripheral presence of internal medicine can be explained by the same fact or, conversely, by the low care coordination role of a significant subgroup of these physicians, given the also prevalent weak longitudinal follow-up profile found in a large proportion of these specialists (Table 8) [2].

Regarding the profile of relationship with authorities, the convergence of authorities in certain medical specialties, in addition to reflecting the prevalent nosology of the patient portfolio, may indicate the concentration of referrals in few professionals considered references or qualified by their peers, and who keep their schedules more widely available for patient appointments [22]. Therefore, in our Amb-HCN, attention should be placed on evaluating access or qualification problems in specialties such as cardiology, endocrinology, gastroenterology, geriatrics, homeopathy, nephrology, neurology, nutrology, pulmonology, psychiatry, and rheumatology. It is also important to note that the fact that some specialties concentrate physicians with a tendency to seek authorities may reflect the intrinsic clinical complexity of their own patient portfolios, a fact supported by the similarity in clinical characteristics and outcomes of patients treated by these two categories (Table 12). In the SNA approach that analyzes networks as mechanisms of social influence, studies suggest that physicians influence and are influenced by the behaviors and practices of colleagues with whom they are in closer contact, leading them to share similar clinical results [9, 21]. It may also be evidence of the tendency of clinical specialists who assume patient care to be knowledgeable about the other specialists that their patients seek [4], giving them greater authority to influence their choices. All of this justifies considering it possible that physicians classified as authorities and those who seek authorities are a cohesive group, with shared patient portfolios and clinical practices.

The third profile proposed in this study aimed to reveal the patterns of patient follow-up by physicians, in light of the assumed responsibility of their specialty. Thus, just as the strong role of patient follow-up by specialists such as pediatrics, internal medicine, geriatrics, psychiatry, endocrinology, nephrology, and others was evident, the weak role of patient follow-up by significant subgroups of physicians in internal medicine, family medicine, and, to a lesser extent, pediatrics was also evident. In the Austrian Amb-HCN, where patient access to physicians was not restricted to primary care physicians as the entry point to the system, Sauter et al. [25] also demonstrated poor performance of family medicine and, to a lesser extent, internal medicine, as coordinators of patient care, judging by the significant proportion of their patients who consulted with other physicians. Efforts should be made to understand to what extent the discrepancy between the practice of these physicians and the standards of their specialty is justified by specific areas of practice [10] or, in turn, explained by the physician’s lack of adherence to expected standards of patient care quality and accessibility.

Another relevant finding of this study is the significant relationship between the centrality, relationship with authorities, and patient follow-up profiles with patient clinical characteristics and outcomes. This relationship may be bidirectional. On the one hand, it suggests that for each patient’s health needs, it is possible and desirable to find the best combination of physician or Amb-HCN profiles that match best with those needs. On the other hand, it suggests that for each physician or Amb-HNC profile, it is possible and desirable to find patients with health needs that best fit those profiles [2, 9]. The objective metrics of Amb-HCN proposed herein can be a valuable aid in identifying the compatibility (or incompatibility) between patients’ health needs and the profile of their physician, allowing healthcare managers to identify service gaps and steer solutions. A study conducted in the Medicare population reported a significant positive association between the number of connections of primary care physicians with other physicians (i.e., degree) and healthcare costs, hospital admissions, days of hospitalization, admissions for primary care-sensitive conditions, emergency department visits, and specialist visits for patients under their care [18]. Another study also found that patients treated by physicians who shared care more intensely with other physicians had higher rates of hospitalization for primary care-sensitive conditions [23]. As in the present study, this relationship can be interpreted as either arising from poor clinical coordination by the primary care physician, or from a greater need for referral to specialists due to the higher clinical complexity of their patients. Although the present study was not designed to evaluate the clinical effectiveness of the network, it is worth mentioning that patients treated by physicians in central positions were those with highest number of chronic comorbidities and yet needed to visit the emergency department or be hospitalized for medical reasons the least (Table 12). Actors in central positions of social networks tend to be considered opinion leaders and highly influential on the clinical decisions of colleagues [2, 9], a fact corroborated in this study, where authorities were more frequently central. The possibility that the physician’s position in the network, their relationship with authorities, and their patient follow-up profile are causally related to patients’ clinical outcomes should be considered by decision-makers and investigated in a timely manner.

The identification of different roles and responsibilities of physicians and specialties supports the theory that the health outcomes of individuals should be attributed not only to individual physicians but also to the functioning of the care network, collaboration, and information flow between physicians and specialties [17, 23, 24]. The demonstration of the natural emergence of self-organized communities of physicians (Table 13 and Box 3), with evident territorial and specialty segregation, reinforces this concept. This finding is a powerful management tool. Landon et al. [5] showed that naturally arranged communities of physicians around territories had professionals with close working relationships and were able to keep most hospitalizations (73%), emergency department visits (40%), primary care visits (88%), and specialists visits (60%) of patients within those networks boundaries. Networks and communities of professionals thus defined would be preferred targets of managers seeking physicians willing to become responsible for the health care of a defined patient population in capitation-based payment contracts [5]. The territorial separation of the communities identified in this study confirms the impression that regionalization is an important attribute of self-constituted Amb-HCN. In this sense, the fact that no medical community with a predominant seat in Contagem municipality - the second most populous municipality in the PHIP coverage area - has emerged, forces us to formulate the hypothesis that beneficiaries living in this municipality may need to seek care in nearby municipalities, such as Belo Horizonte or Betim, probably due to the insufficient specialty network in Contagem.

Several strengths of this study can be highlighted. By using administrative claim data routinely collected by the PHIP and by considering all more than 1 million beneficiaries of a healthcare plan which does not restrict the location or physician for consultations, the study has no missing data, avoided selection and response bias, and can be considered representative of a large population. By relying on the date of consultation, it was possible to model the network as directional, which has been quite uncommon in published studies, in which the relationship between physicians has generally been treated as non-directional.

Some limitations of this study should also be acknowledged. As with any quantitative and cross-sectional representation of reality, the application of SNA certainly cannot capture all the complexity involved in the emerging relationships between actors. Part of the structural and functional topology observed in our Amb-HCN may originate from conjunctural, unstable, or seasonal factors that were not considered in the analysis. In addition, SNA requires multiple methodological choices appropriate for the study objectives but not necessarily relevant for all other purposes [2, 10]. Therefore, the extent to which the findings are reproducible and stable over time is unknown. Another limitation inherent to SNA is the influence of actors beyond the boundaries of the analyzed network. If the analysis did not include all relevant actors, it is unlikely that the results captured all the complexity of the Amb-HCN. Thirdly, as the analysis used administrative data and may not necessarily represent explicit and deliberate referrals between physicians, some of the relationships found may be spurious. An ideal approach to this problem, although methodologically complex, would be to restrict physician relationships to episodes of care or specific health problems of the patients, which would exclude circumstantial relationships between physicians who treat patients for unrelated health problems. This was the reason why this study considered consultations spaced by an interval between 7 and 45 days. Finally, although the analysis focused on the role of physicians as the main actors, it would be interesting to know to what extent the structure of the Amb-HCN depends on patient behaviors and preferences.

## Conclusions

Viewing our Amb-HCN as a social network and applying measures based on graph theory and SNA provided emerging insights into the most influential actors and specialties, potential gaps in care, and the most prevalent diseases in our patient portfolio. The identification of self-constituted Amb-HCN can form a rational basis for developing more formal networks or monitoring patient care performance without assigning responsibility to a single physician. However, transferring research knowledge into actionable plans and decision-making by health authorities requires reflection, business expertise, and strategies based on continuous improvement cycles. The way network metrics reflect attributes of quality, access, and care coordination in healthcare is an evolving field [9]. Defining operational metrics for the roles and responsibilities of healthcare professionals, understanding the functional structure of Amb-HCN, and evaluating their influence on patient health outcomes remain challenges for researchers and health policy makers.

## Data Availability

All relevant data are within the manuscript and its Supporting Information files.

## Supporting information

**S1 Table. Distribution of physicians according to consultation productivity and medical specialty**

**S2 Table. Pearson correlation coefficients between vertex-level measures**

